# Fully automatic segmentation of craniomaxillofacial CT scans for computer-assisted orthognathic surgery planning using the nnU-Net framework

**DOI:** 10.1101/2021.07.22.21260825

**Authors:** Gauthier Dot, Thomas Schouman, Guillaume Dubois, Philippe Rouch, Laurent Gajny

## Abstract

**Objectives:** To evaluate the performance of the nnU-Net open-source deep learning framework for automatic multi-task segmentation of craniomaxillofacial (CMF) structures in CT scans obtained for computer-assisted orthognathic surgery.

**Methods:** Four hundred and fifty-three consecutive patients having undergone high-resolution CT scans before orthognathic surgery were randomly distributed among a training/validation cohort (*n* = 300) and a testing cohort (*n* = 153). The ground truth segmentations were generated by 2 operators following an industry-certified procedure for use in computer-assisted surgical planning and personalized implant manufacturing. Model performance was assessed by comparing model predictions with ground truth segmentations. Examination of 45 CT scans by an industry expert provided additional evaluation. The model’s generalizability was tested on a publicly available dataset of 10 CT scans with ground truth segmentations of the mandible.

**Results:** In the test cohort, mean volumetric Dice Similarity Coefficient (vDSC) & surface Dice Similarity Coefficient at 1mm (sDSC) were 0.96 & 0.97 for the upper skull, 0.94 & 0.98 for the mandible, 0.95 & 0.99 for the upper teeth, 0.94 & 0.99 for the lower teeth and 0.82 & 0.98 for the mandibular canal. Industry expert segmentation approval rates were 93% for the mandible, 89% for the mandibular canal, 82% for the upper skull, 69% for the upper teeth and 58% for the lower teeth.

**Conclusion:** While additional efforts are required for the segmentation of dental apices, our results demonstrated the model’s reliability in terms of fully automatic segmentation of preoperative orthognathic CT scans.

**Key points:**

- The nnU-Net deep learning framework can be trained out-of-the-box to provide robust fully automatic multi-task segmentation of CT scans performed for computer-assisted orthognathic surgery planning.
- The clinical viability of the trained nnU-Net model is shown on a challenging test dataset of 153 CT scans randomly selected from clinical practice, showing metallic artifacts and diverse anatomical deformities.
- Commonly used biomedical segmentation evaluation metrics (volumetric and surface Dice Similarity Coefficient) do not always match industry expert evaluation in the case of more demanding clinical applications.

## Introduction

Orthognathic surgery addresses congenital and acquired conditions of the facial skeleton by repositioning the jaws into a functional relationship in subjects presenting dentofacial deformities. It has been reported that up to 5% of the USA and UK populations could require orthognathic surgery [1]. Surgical correction of craniomaxillofacial (CMF) deformities requires defining a specific surgical treatment plan for every single patient [2]. In recent years, several teams have shown the reliability of computerized methods to analyze dentofacial deformities or to elaborate surgical treatment plans [2, 3]. Virtual planning is usually performed using a CT scan or cone-beam CT (CBCT) scan of the patient’s head [3]. The first step in the planning pipeline involves the extraction of structures of interest from the CT data by semantic segmentation. The choice of (CB)CT scan resolution, field of view (FOV) and anatomical structures to be segmented depend on what the planning is intended for. For the purpose of surgical guides or personalized implant manufacturing, the orthognathic surgery planning is based on high-resolution CT scans (with voxel size around 0.5*0.5*0.5mm^3^) with full-head FOV (around 250mm). The following anatomical structures need to be segmented: upper skull and mandible, including the mandibular canals, upper and lower teeth (crowns and roots). Proper segmentation and delineation of these structures is known to be challenging due to factors such as large interindividual morphological variations, tight connections between the structures, lack of contrast in joints and teeth apices, frequent presence of artifacts (orthodontic materials, fixation implants, dental fillings or crowns) [4, 5].

Semi-automatic algorithms may be used to make segmentation less time-consuming [6]. Some of these methods provide high segmentation accuracy, yet are not fully automatic and therefore still require time-consuming manipulations by trained operators. In recent years, segmentations of medical images using deep learning algorithms have outperformed previously used algorithms. Deep learning algorithms might indeed be able to perform fully automatic segmentations [7]. Several authors have developed specific deep learning-based models for automatic segmentation of the upper skull, mandibular bone, teeth or mandibular canal [5, 8–18]. Most of these approaches relied on a U-Net convolutional architecture [19], and yielded promising results, with reported volumetric Dice Similarity Coefficient (vDSC) between 90% and 95%. However, some limitations restrict their clinical applicability. None of them specified whether the dataset used for training and testing the algorithm included routine clinical cases, nor how the scans were selected. When the evaluation of the model on a hold-out test dataset was provided, the number of test scans was always less than 30. Moreover, none of the algorithms in the studies were used to segment all the structures of interest for computer-assisted planning of orthognathic surgery and personalized implant manufacturing, and only one work segmented bone and teeth separately [18]. This calls into question the reproducibility and generalizability of previously published results, which are crucial factors for clinical validity [7, 20].

Recently, the nnU-Net framework was proposed as an out-of-the-box tool which automatically configures itself in order to perform deep learning-based biomedical image segmentation [21]. This tool is publicly available and was shown to surpass most existing models on 23 public datasets used in international biomedical segmentation competitions. nnU-Net could be helpful for direct clinical applications, as it is open source and does not necessarily require expert knowledge to obtain competitive results. To our knowledge, the performance of this tool has not yet been evaluated for the segmentation of craniofacial hard tissue in a clinical context.

In this work, our main objective was to evaluate the performance of the nnU-Net framework for automatic segmentation of CMF structures in routine CT scans performed for computer-assisted orthognathic surgery.

## Materials and methods

### Patient selection

Data were selected from a retrospective cohort of all consecutive patients having undergone orthognathic surgery in a single maxillofacial surgery department between January 2017 and December 2019. Patients referred to this center presented a wide variety of dentofacial deformities, came from various socioeconomic backgrounds and were ethnically diverse. Patients were considered for inclusion whatever dental deformity they presented, with no minimum age. Exclusion criteria were refusal to participate in the research (all patients were contacted by mail) and lack of industry-certified CT scan segmentations (see next paragraph: “Ground truth segmentation process”). Of the 473 subjects who underwent orthognathic surgery within the study timeframe, 4 refused to participate and 16 lacked industry-certified CT scan segmentations. 453 subjects (453 CT scans) were eventually included in our dataset (Fig. 1). Mean in-plane pixel size of the CT scans was 0.45*0.45mm^2^ and their mean slice thickness was 0.34mm. Most CT scans (*n* = 417) were obtained using a GE Healthcare Discovery (GEHC) CT750HD scanner with a tube current of 50mA, an exposure time of 730s and a tube voltage of 100kV. Scans were randomly distributed among a train/validation set (*n* = 300) and test set (*n* = 153). CT scans characteristics and CT machines are detailed in Table 1. This study was ethically approved by an Institutional Review Board.

**Table 1.** CT scan characteristics and CT machines in the train/validation, test and public mandible test datasets.

|  | Train/Validation | Test | Public Mandible Test [22] |
| --- | --- | --- | --- |
| Number of CT scans | 300 | 153 | 10 |
| Mean in-plane pixel size (mm<sup>2</sup>) | 0.44 * 0.44 | 0.45 * 0.45 | 0.45 * 0.45 |
| Mean slice thickness (mm) | 0.33 | 0.34 | 0.6 |
| Number of scans by CT Machine |  |  |  |
| GEHC Discovery CT750 HD | 284 | 144 |  |
| GEHC Optima CT540 or CT660 | 7 | 5 |  |
| Siemens Sensation 64 |  |  | 10 |
| Other CT Machine<sup>1</sup> | 9 | 4 |  |

GEHC: GE Healthcare. <sup>1</sup>GEHC Revolution CT, Philips Ingenuity Core, Siemens SOMATOM Definition AS, Toshiba Aquilion Prime SP

**Figure 1.**
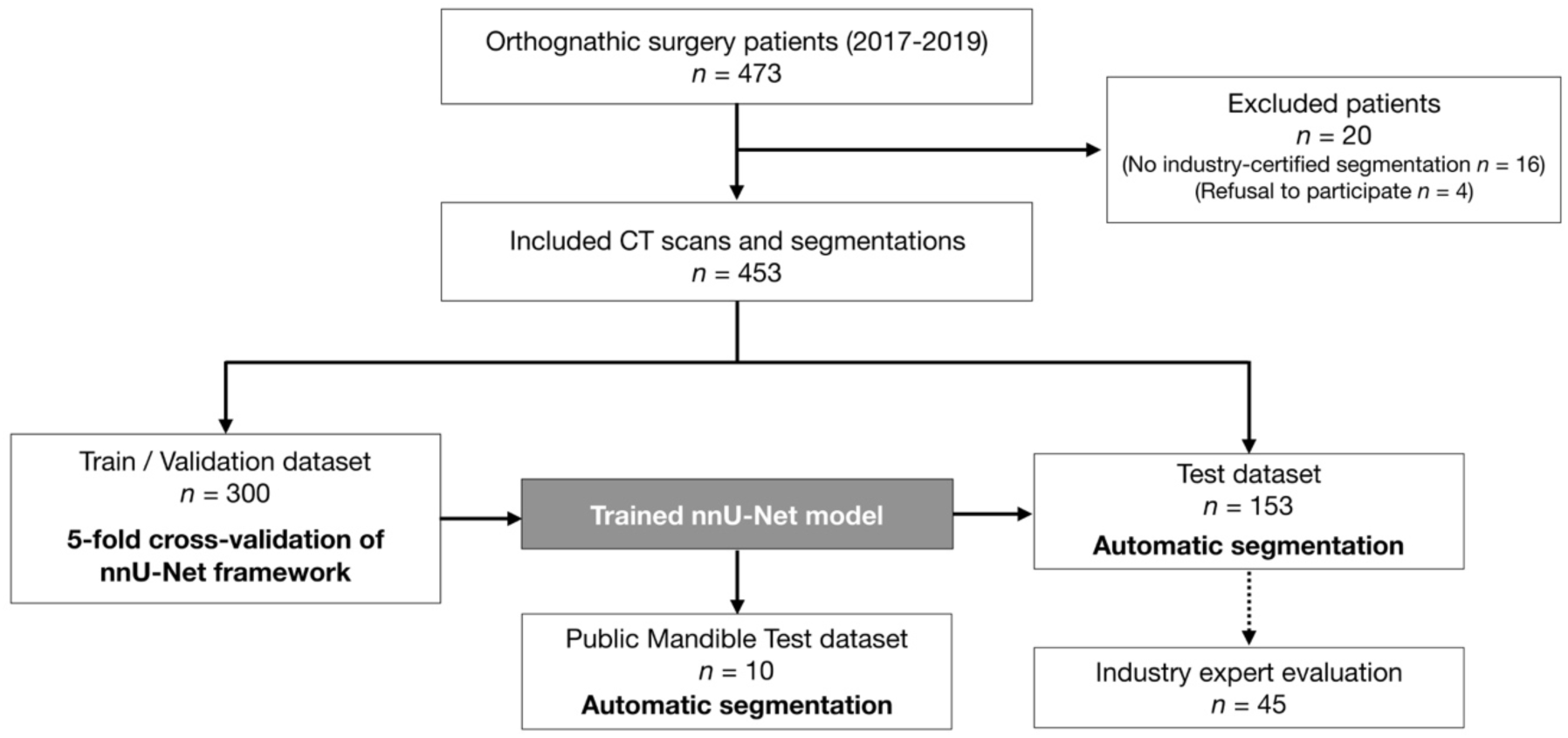
Data flow of the patient selection, training and evaluation process.

### Ground truth segmentation process

All CT scans had been segmented prior to our study during patient treatment. Ground truth segmentations were used for diagnosis, computer-aided surgical planning and manufacturing of 3D-printed personalized surgical guides and fixation implants according to a certified internal procedure (Materialise). This industry procedure is confidential and cannot be fully described here. It implies semi-automatic segmentations, manually refined by a first operator [Step 1] before slice-by-slice verification for validation by a senior operator [Step 2] focusing mainly on the regions of interest: external surface of the bones, teeth and mandibular canals. Steps 1 & 2 are repeated until the segmentations are approved and certified for clinical use. This process results in 5 segmentation masks: (1) upper skull, (2) mandible, (3) upper teeth, (4) lower teeth and (5) both mandibular canals.

### Public mandible test dataset

In order to assess the generalizability of our trained model, we tested it on a public dataset of 10 high-resolution CT scans from clinical practice [22]. These scans had the particularity of presenting complete mandibular bone structures entirely devoid of teeth. Ground truth manual segmentations of the mandible by two experts (A and B) were provided, which differed from our segmentations in that they were filled. Table 1 provides more details about this dataset. To the best of our knowledge, this is the only publicly available dataset of high-resolution Head CT scans with ground truth segmentation of one of the structures of interest for orthognathic surgery planning.

### nnU-Net framework

The nnU-Net deep learning framework was used as an out-of-the-box tool, following instructions given by Isensee *et al*. [21]. Our raw train/validation dataset was used to automatically configure preprocessing, network architecture, training and post-processing pipelines. No modifications were made in setting the nnU-Net hyperparameters and data augmentation strategy. The training of 3D full resolution U-Net was performed on our train/validation set according to a 5-fold cross-validation strategy. After the end of the training pipeline, cross-validation result analysis showed that the model incorrectly labeled a few voxels as teeth in some scans displaying no upper and/or lower teeth. As a result, we implemented additional post-processing for teeth masks, consisting in removing all components smaller than an empirically-determined threshold. Finally, inference was performed on our test dataset as well as on the public mandible dataset (Fig. 1). More details on the implementation of the deep learning framework are provided in Supplementary Information.

### Evaluation metrics

Quantitative evaluation of the model performance was performed on our test set by comparing ground truth masks with predictions for each of the 5 segmentation masks. We followed best practice in evaluating model results, using both volume-based and surface-based metrics. Our main volume-based metric was the commonly used vDSC. Our main surface-based metric was surface DSC at 1mm (sDSC). Compared with classical metrics such as vDSC, sDSC, which was introduced recently, has been shown to be more strongly correlated with the amount of time needed to correct a segmentation for clinical use [7]. We set our acceptable tolerance of sDSC at 1mm, as was done in recent international challenges in biomedical imaging. As in previous studies, an sDSC score of 95% was chosen as threshold value to consider variations between two segmentations as clinically non-significant. Additional quantitative metrics were computed after the common biomedical segmentation evaluations: Jaccard Coefficient, Volumetric Similarity, Average Surface Distance and Hausdorff distance.

### Industry expert evaluation procedure

A random sample of our predicted masks for 45 subjects from our test dataset was sent to industry experts (Materialise) to be evaluated according to the 2-step validation process described above. The experts were blinded to our results and did not know that these segmentations were automatic, as they were evaluated among the flow of “classical” segmentations. Each segmentation mask was labeled “validated for clinical use” or “not validated for clinical use”.

### Statistical analysis

Continuous variables are presented as mean ± standard deviation and categorical variables are expressed as numbers and percentages. vDSC and sDSC results are presented as percentages (%). The Wilcoxon test was used to compare vDSC and sDSC in scans obtained with GEHC CT750HD and scans obtained with other machines; p-values < 0.05 were considered significant. All data were analysed with Python (v.3.7) and RStudio software (v.1.3).

## Results

### Patient characteristics

In our database, patient mean age was 27 ± 11 years (minimum age 14, maximum age 66). The patients presented diverse anatomical deformities. 237 subjects (52.3%) exhibited skeletal class II (prognathic maxilla and/or retrognathic mandible), 163 subjects (36.0%) exhibited skeletal class III (retrognathic maxilla and/or prognathic mandible) and 165 subjects (36.4%) displayed asymmetry (clear asymmetry of the maxilla and/or the mandible evaluated on the 3D models). 89.6% of the CT scans (*n =* 406) showed metallic artefacts, in the form of orthodontic materials for 354 subjects (77.9%), metallic dental fillings or crowns for 193 subjects (42.6%) and fixation implants from previous orthognathic surgeries for 30 subjects (6.6%). Some subjects had no upper teeth (*n =* 4), no lower teeth (*n =* 1) or no teeth at all (*n =* 2). The public mandible test dataset included senior patients (63 ± 9 years) with no teeth at all. Table 2 summarizes patient characteristics in our datasets.

**Table 2.** Descriptive characteristics of the patients in the train/validation, test and public mandible test datasets.

| Characteristic | Train/Validation ($n = 300$) | Test ($n = 153$) | Public Mandible Test [22] ($n = 10$) |
| --- | --- | --- | --- |
| Age, mean ± SD, years | 27 ± 11 | 26 ± 9 | 63 ± 9 |
| Gender, no. (%) |  |  |  |
| Female | 178 (59.3) | 83 (54.2) | 5 (50.0) |
| Male | 122 (40.7) | 70 (45.8) | 5 (50.0) |
| Skeletal deformity, no. (%) |  |  |  |
| Class I | 35 (11.7) | 18 (11.8) | X<sup>e</sup> |
| Class II<sup>a</sup> | 154 (51.3) | 83 (54.2) | X<sup>e</sup> |
| Class III<sup>b</sup> | 111 (37.0) | 52 (34.0) | X<sup>e</sup> |
| Asymmetry<sup>c</sup> | 112 (37.3) | 53 (34.6) | 1 (10.0) |
| Syndromic deformity | 8 (2.7) | 8 (5.2) | X<sup>f</sup> |
| Absence of teeth, no. (%) |  |  |  |
| No upper teeth | 4 (1.3) |  |  |
| No lower teeth | 1 (0.3) |  |  |
| No teeth at all | 1 (0.3) | 1 (0.7) | 10 (100) |
| Metal artifacts, no. (%) |  |  |  |
| Orthodontic materials | 232 (77.3) | 122 (79.7) |  |
| Metallic dental filling/crown | 126 (42.0) | 67 (43.8) |  |
| Fixation implants<sup>d</sup> | 17 (5.7) | 13 (8.5) |  |
| No metallic artifact | 35 (11.7) | 12 (7.8) | 10 (100) |

<sup>a</sup>Prognathic maxilla and/or retrognathic mandible. <sup>b</sup>Retrognathic maxilla and/or prognathic mandible. <sup>c</sup>Clear asymmetry of the maxilla and/or the mandible evaluated on the 3D models. <sup>d</sup>Fixation implants from a previous surgery. <sup>e</sup>Skeletal classification cannot be provided due to missing teeth and lack of information about the mandible's vertical position during CT scan acquisition. <sup>f</sup>No information provided with the database. SD, Standard Deviation.

### Model performance

#### Quantitative evaluation on our test dataset

The mean results of vDSC and sDSC for each segmentation label of our test set are shown in Table 3, while Figure 2 shows the distribution of the results. The mean vDSC for all masks was 92.24 ± 6.19%. Without the mandibular canal results, the mean vDSC was 94.90 ± 0.91%. The mean sDSC for all masks was 98.03 ± 2.48%. Out of the 153 scans, 148 presented a mean sDSC for all masks which cleared the 95% limit for clinical significance. There were no statistically significant differences in both vDSC and sDSC when comparing scans obtained with GEHC CT750HD and scans obtained with other machines. Additional quantitative evaluation (Jaccard Coefficient, Volumetric Similarity, Average Surface Distance and Hausdorff distance) results for cross-validation and test datasets are provided in Supplementary Materials.

**Table 3.** Mean vDSC and sDSC results on our test dataset (*n* = 153).

| Metric, mean ± SD [%] | Upper Skull | Mandible | Upper teeth | Lower Teeth | Mandibular canal | Total |
| --- | --- | --- | --- | --- | --- | --- |
| vDSC | 96.22 ± 1.43 | 94.19 ± 1.62 | 94.83 ± 1.81 | 94.38 ± 2.32 | 81.59 ± 5.79 | 92.24 ± 6.19 |
| sDSC | 96.92 ± 3.08 | 97.92 ± 1.22 | 98.87 ± 1.18 | 98.53 ± 2.00 | 97.9 ± 3.51 | 98.03 ± 2.48 |

**Figure 2.**
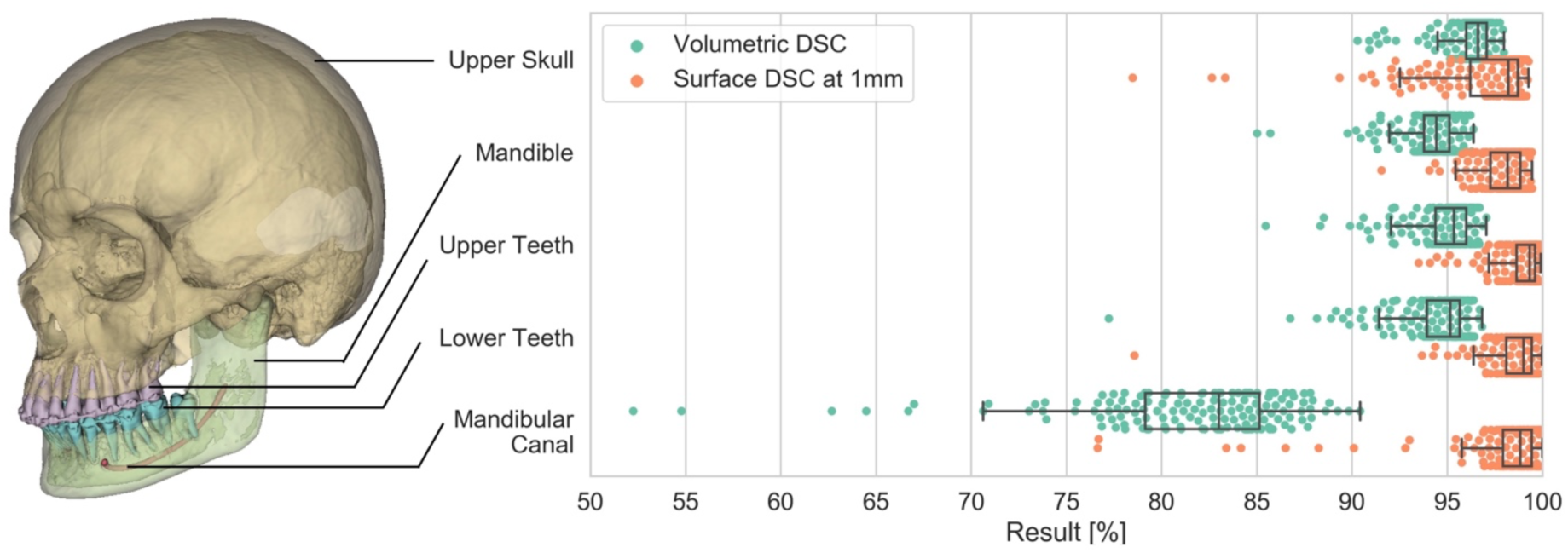
Left side: 3D model reconstructed from predicted segmentation masks (Upper Skull and Mandible with transparent overlay). Right side: distribution of vDSC and sDSC results in our test dataset (153 CT scans), for each segmentation mask (no result below 50%).

Four subjects representative of our test dataset and of the anatomic diversity of the database were chosen to illustrate our results (Fig. 3, Fig. 4, Supplementary videoclips 1 to 4). The most notable segmentation error made by the model on these subjects was the incorrect labeling of an upper deciduous tooth as a lower tooth in a patient suffering from craniofacial syndrome with several impacted teeth (Fig. 3b, Fig. 4b and Supplementary videoclip 2).

**Figure 3.**
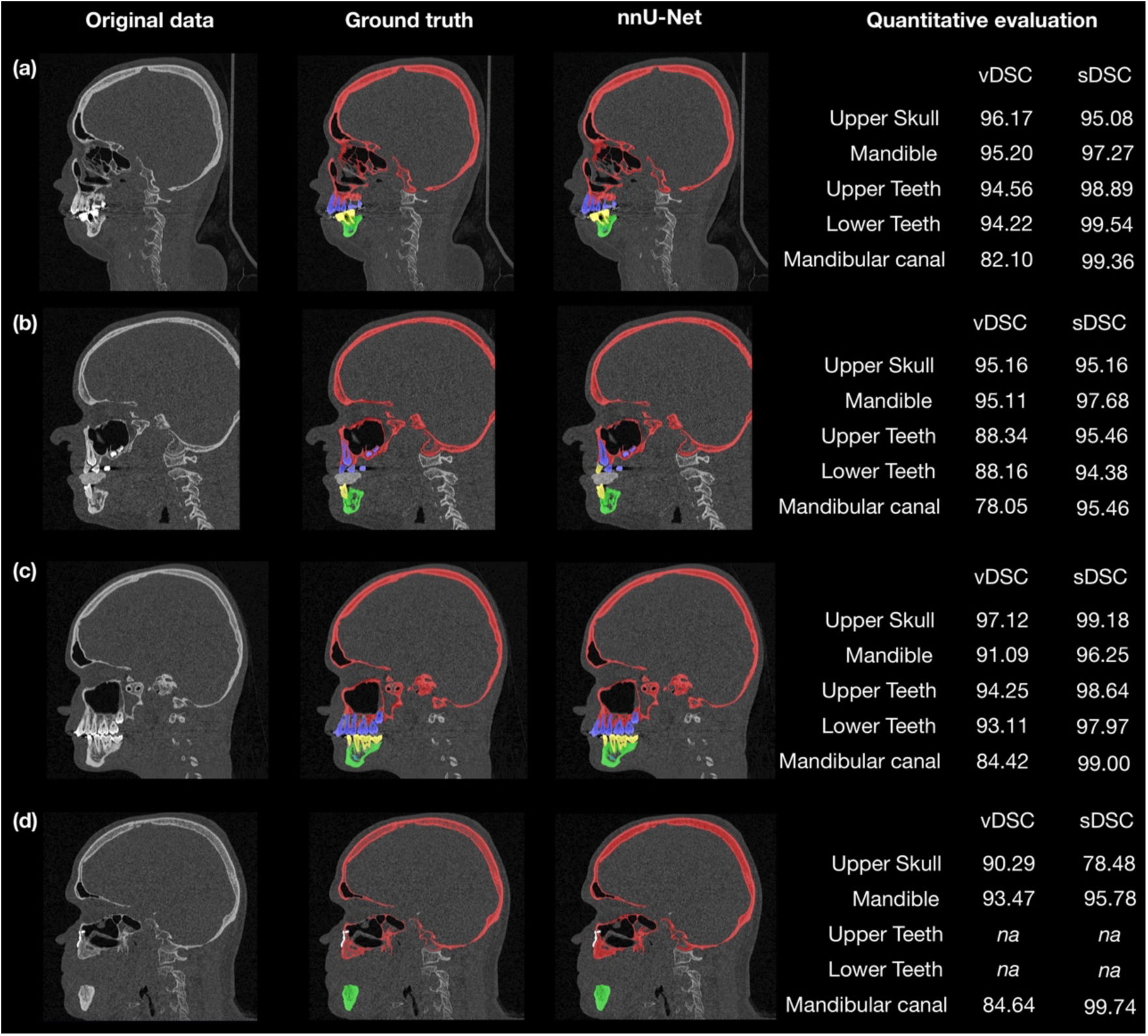
Four representative cases (a to d) showing sagittal slices of original data, ground truth segmentations, nnU-Net network-predicted segmentations and quantitative evaluation results. Red, upper skull; green, mandible; blue, upper teeth; yellow, lower teeth; cyan, mandibular canal.

**Figure 4.**
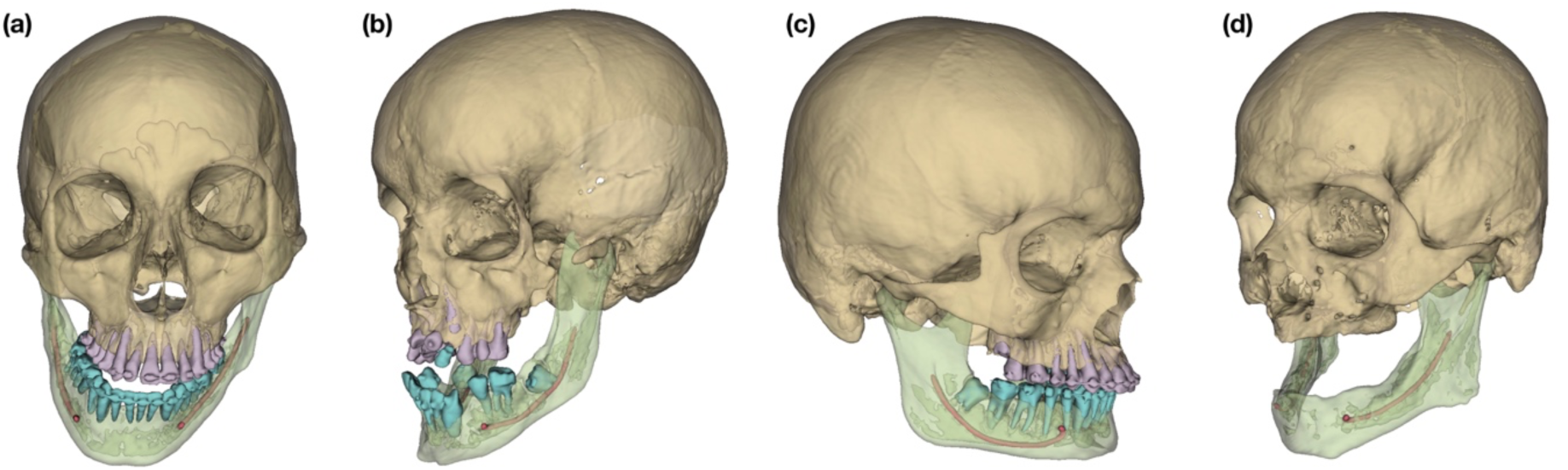
3D surface models of segmentation results for 4 subjects representative of the anatomical diversity and of the challenges arising from our test dataset: (a) prognathic and asymmetric mandible; (b) craniofacial syndrome, with included and missing teeth; (c) retrognathic mandible; (d) no teeth and maxillary fixation implants from previous surgery (not segmented by the network).

#### Expert evaluation on our test dataset

The results of the industry expert validation process are shown in Table 4. Validation rates were about 90% for mandible and mandibular canals, 80% for upper skull and 60% for teeth. In total, 19 (42.2%) CT scans had all their segmentation masks validated. When excluding dental masks, 34 (75.6%) CT scans had their segmentation masks validated. Three CT scans out of the 45 (6.7%) did not show any metallic artefact, and had their 5 segmentation masks validated by industry experts. Comments appended to non-validated cases for bones mentioned small holes on the bony surface or under-segmentation of the anterior nasal spine. Reasons for rejection of mandibular canal masks were the inclusion of a few outlier voxels or the omission of an auxiliary canal during segmentation. As to teeth, reasons for non-validation were under- or over-segmentation of a few voxels of the apex (Fig. 5).

**Table 4.** Results of industry expert evaluation on 45 random CT scans from our test set.

| Result, no. (%) | Upper Skull | Mandible | Upper teeth | Lower Teeth | Mandibular canal | Total | Total without teeth masks |
| --- | --- | --- | --- | --- | --- | --- | --- |
| Validated | 37 (82.2) | 42 (93.3) | 31 (68.9) | 26 (57.8) | 40 (88.9) | 19 (42.2) | 34 (75.6) |
| Not validated | 8 (17.8) | 3 (6.7) | 14 (31.1) | 19 (42.2) | 5 (11.1) | 26 (57.8) | 11 (24.4) |

**Figure 5.**
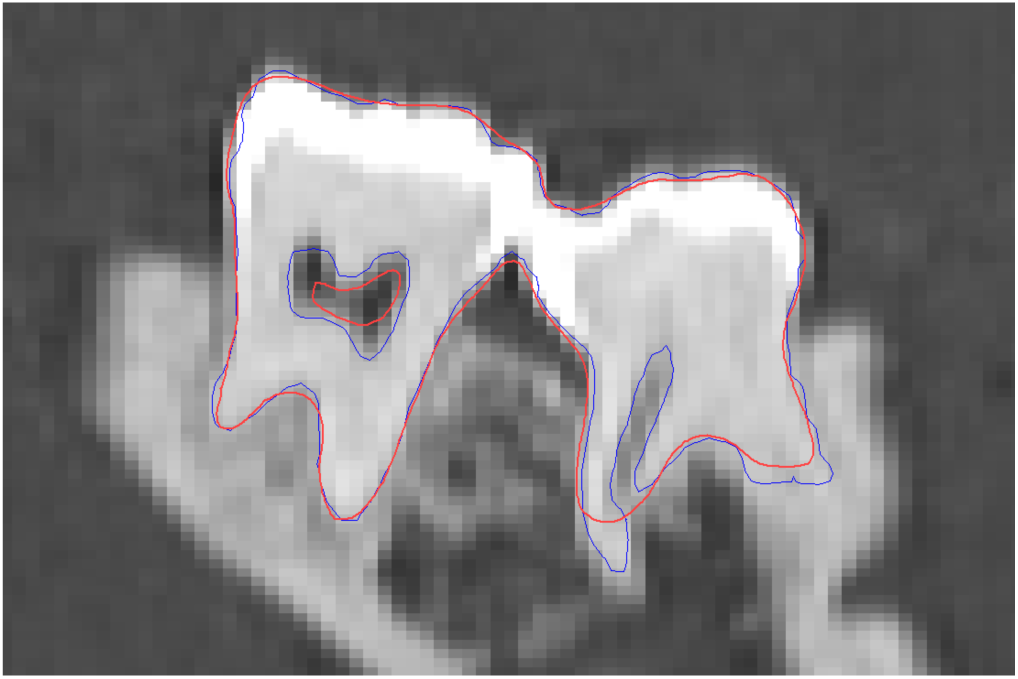
Representative lower teeth mask which was not validated by industry expert. Red line: predicted mask contour. Blue line: ground truth mask contour.

#### Quantitative evaluation on public mandible dataset

vDSC and sDSC results of the inference on the public mandible dataset are provided in Table 5. Most of the model’s errors were located in the anterior part of the mandible, where the edentulous patients’ alveolar bone was atrophic and extremely thin (Fig. 6). One scan (mandible #10), performed on a subject with an endotracheal tube, produced low-quality results. Additional quantitative evaluation results for this dataset are presented in Supplementary Table 3.

**Table 5.**
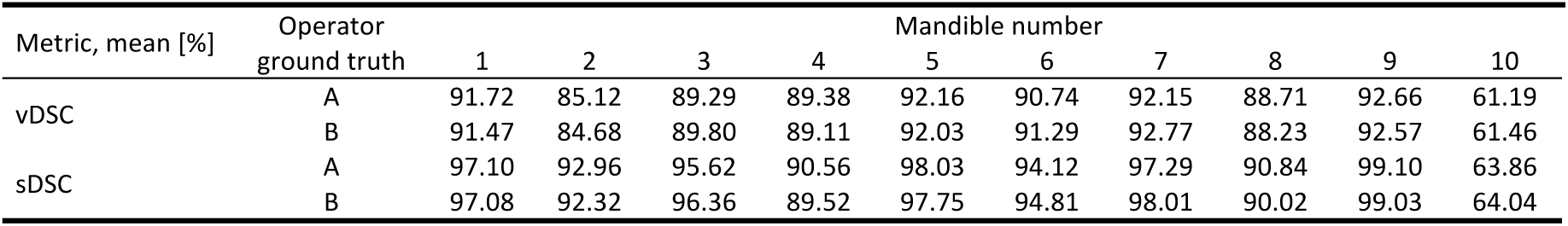
vDSC and sDSC results on public mandible dataset.

**Figure 6.**
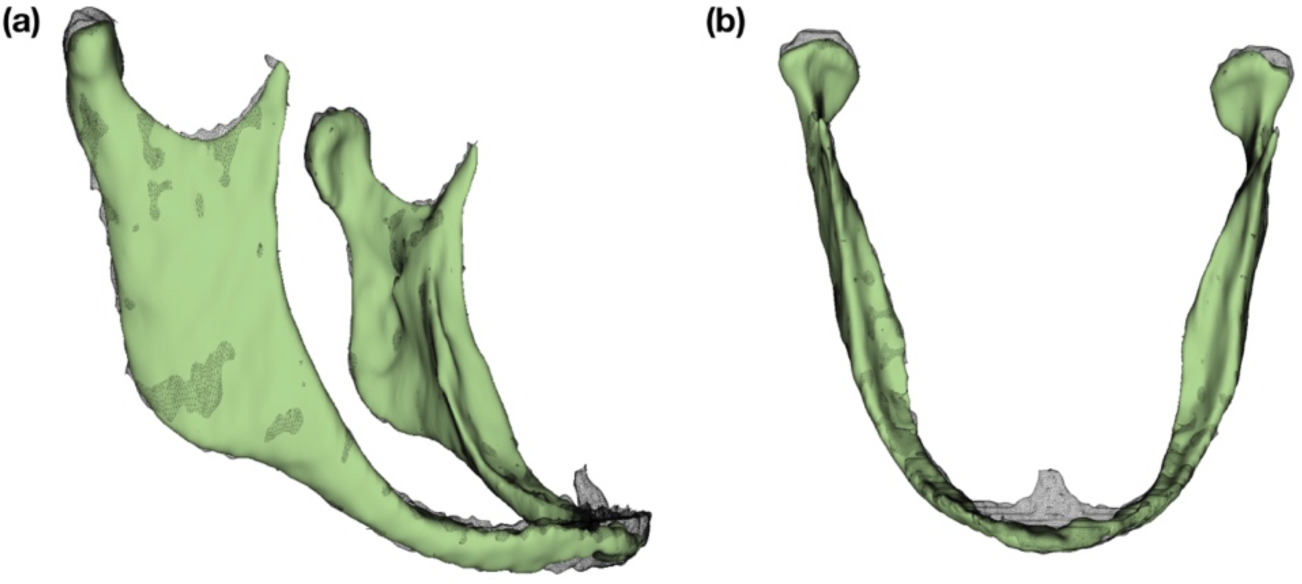
3D surface models of ground truth and automatic segmentation for mandible #2 of public mandible test dataset: black wireframe, ground truth (operator A); solid green, automatic segmentation result. (a) right lateral view; (b) upper view.

### Training and automatic segmentation times

Training time for one fold on one GPU was about 48 hours (1,000 epochs). The trained model provided automatic segmentation of 1 CT scan in approximately 10 minutes.

## Discussion

The main goal of this study was to evaluate the performance of the nnU-Net framework for semantic segmentation of CMF CT scans obtained for the planning of orthognathic surgeries. We chose this deep learning framework because it is open source and well-maintained, and has the ability to automatically configure itself without manual intervention. It has delivered state-of-the art segmentation results on a large diversity of biomedical datasets, surpassing most highly-specialized algorithms [21]. Our quantitative results demonstrated the model’s relevance for CT scan segmentation, with 97% of our test dataset showing a mean sDSC above 95%. However, this study also illustrated the challenges arising from the evaluation of deep learning-based algorithms in the case of demanding, specific clinical applications which forbid blind trust in quantitative metrics [7, 23]. For example, our sDSC results for upper skull masks showed a relatively large dispersion, which is not reflected, however, in expert evaluation results. Indeed, discrepancies between ground truth and prediction masks were mainly found in small bony structures located inside the skull, which are not relevant for most clinical applications (Fig. 3d). Conversely, the number of non-validated teeth segmentation masks cannot be directly correlated to quantitative results, most masks obtaining excellent vDSC and sDSC results (>95%). No teeth labels were rejected based on segmentation errors localized at crown level, despite the difficulty in delineating the upper and lower teeth when they are in contact or show metallic artifacts [9, 17]. Instead, they were rejected because of a few mislabeled voxels located in zones (root apices) clinically relevant for personalized implant manufacturing. However, many other clinical applications, such as computer-assisted diagnosis or planning not involving personalized implant manufacturing, would not require such precision in the segmentation of dental apices. Finally, the clinical value of sDSC metrics in the context of small object segmentation was demonstrated by our mandibular canal segmentation results, for which vDSC did not seem like an appropriate metric [7, 23].

This study, which followed best practice guidelines [20], is the first to train and test a deep learning segmentation model on such a large number of high-resolution CMF CT scans. Our results are comparable or superior to those of previously published studies, despite the more challenging task we faced: all previous results were based on smaller test datasets (between 0 and 30 scans) and fewer segmentation masks [5, 8–18]. No previous work had included a cohort of consecutive patients, and only one previous publication had clearly stated that its database included patients with syndromic conditions [16]. Our results for the segmentation of scans from patients with marked syndromic deformities show the versatility of the model (Fig. 3b), and are comparable to those of Wang *et al*. who recently reported on a deep learning-based model for multi-task segmentation of bone and teeth separately [18]. However, the latter study lacked a hold-out test dataset, included only scans devoid of metal artifacts and did not differentiate between maxillary and mandibular structures. Our results for the segmentation of mandibular canals (vDSC of 81.59% ± 5.79%, sDSC of 97.9% ± 3.51%) are significantly superior to those reported by Jaskari *et al*. (vDSC of 57%) and Kwak *et al*. (average Jaccard coefficient of canal and background of 57.72%) [10, 11]. However, most existing studies used CBCTs, which are more difficult to segment than CT scans [16]. In addition, the absence of a public dataset of full high-resolution CMF (CB)CT scans with ground truth segmentations prevents direct comparison of our trained nnU-Net model with previously described ones.

This research has several limitations. It is a single-center study, and thus cannot assess the reliability of the results in other cohorts. Generalization of our results to other CT machines or deformities was partially evaluated on 10 public CT scans obtained using a different CT machine and presenting very different anatomies from those in our training dataset. No subjects from our training database had such edentulous mandibles or endotracheal tube. Setting aside mandible #10 on the basis of endotracheal tube interference, our vDSC results nonetheless outperformed semi-automatic open source segmentation algorithms tested on the same dataset [6]. This suggests that the trained nnU-Net model presented in this study could be used in other settings and for other clinical applications in the fields of CMF or dentistry requiring high-resolution CT scans, although proper multicenter and prospective evaluations are still needed. In order to assess the efficacy of this trained nnU-Net model in clinical practice, we plan on evaluating its use prospectively for orthognathic surgery planning and personalized implant manufacturing. As our deep learning model was trained with a target size of 0.31*0.45*0.45mm^3^, it is not expected to provide competitive results on CT scans with much lower resolutions (resolutions around 2.5*1*1mm^3^ used in other clinical contexts [7], for example). Our study focused on CT scans because it is the imaging modality we use for computer-assisted planning of orthognathic surgery at this time. In future works we plan to fine-tune this model in order to evaluate its performance with CBCT scans, another widespread and challenging imaging modality. Finally, we will attempt to use our segmentation results for automatic localization of anatomic landmarks, in order to provide cephalometric measurements for clinical diagnosis and treatment planning [12, 16, 17, 24].

To conclude, this study showed that nnU-Net could be used out-of-the-box (along with a simple post-processing volume filter) to provide robust segmentation of routine preoperative CMF CT scans. While the successful segmentation of dental apices will require additional efforts, quantitative results and industry expert evaluation demonstrated the clinical validity of our trained model for the segmentation phase of computer-assisted orthognathic surgery planning. Our results suggest that the nnU-Net framework could be trained from scratch easily, using databases from other departments, to answer the specific needs of many clinical setting.

## Data Availability

Some study subjects (10 mandibular CT scans) were shared in a previous publication: Wallner J, Mischak I, Jan Egger (2019) Computed tomography data collection of the complete human mandible and valid clinical ground truth models. Sci Data 6:190003. https://doi.org/10.1038/sdata.2019.3

## Abbreviations

CBCT: Cone Beam CT
CMF: Craniomaxillofacial
FOV: Field of view
sDSC: Surface Dice Similarity Coefficient at 1mm
vDSC: Volumetric Dice Similarity Coefficient

## Acknowledgment

The authors would like to thank Fabian Isensee and all the team behind the nnU-Net framework for sharing their great work, and the “Association Les Chirurgiens Maxillo-Faciaux” for its technical support.

## Compliance with Ethical Standards

### Guarantor

The scientific guarantor of this publication is Laurent Gajny.

### Statistics and Biometry

One of the authors has significant statistical expertise.

### Informed Consent

Written informed consent was waived by the Institutional Review Board. All patients were contacted by mail and were offered to refuse to participate in the research.

### Ethical Approval

The IRB “Comité d’Ethique pour la Recherche en Imagerie Médicale” (CERIM) gave ethical approval for this research (number CRM-2001-051).

### Methodology

Methodology:

- retrospective
- diagnostic or prognostic study
- performed at one institution

