## Supplementary Information for "Fully automatic segmentation of craniomaxillofacial CT scans for computer-assisted orthognathic surgery planning using the nnU-Net framework"

### Implementation Details.

All experiments were performed using the nnU-Net v.1.6.5 framework running on an Nvidia Pytorch Docker container (v.20.10-py3) on our laboratory workstation (CPU AMD Ryzen 9 3900X 12-Core; 128 Gb RAM; GPU Nvidia Titan RTX 24Gb). Our preliminary tests showed that the 3D U-Net full-resolution model far outperformed the 2D U-Net model, while 3D U-Net cascade performances were inconsistent. As a result, we focused on training the 3D U-Net full-resolution model. Target spacing of the model was 0.31*0.45*0.45mm, patch size was 192*112*112 pixels and batch size was set to 2.

Training was performed once on our train/validation set following a 5-fold cross-validation strategy. Training time for one fold was about 48 hours (1,000 epochs) and the GPU VRAM memory footprint was about 8GB. Our automatic post-processing strategy showed that removing all but the largest components improved performance in segmentation masks of upper skull and mandible. After the end of the training pipeline, we assessed the need for additional post-processing by looking at the predictions with the worst results. Analysis of cross-validation results showed that the model incorrectly labeled a few voxels as teeth in some scans displaying no upper and/or lower teeth. As a result, we implemented further post-processing for teeth masks, using SimpleITK library to remove all components smaller than a threshold which we empirically set at $60mm^3$, i.e. the largest volume that improved cross-validation results. Cross-validation quantitative results (300 CT scans) are provided in Supplementary Table 1 and Supplementary Figure 1. Inference was performed once with TTA, then once without, on the test datasets. Inference took approximately 45 minutes per CT scan with TTA, compared to 10 minutes without. Since disabling TTA did not negatively affect the prediction results, we chose to present results for inference without TTA. The post-processing strategy described above was applied to the prediction results.

Our quantitative metrics were computed using SimpleITK library v.2.0.2 (for vDSC, Jaccard Coefficient, Volumetric Similarity, Average Surface Distance and Hausdorff distance) and Medical Segmentation Decathlon Challenge implementation (for sDSC).

For the public mandible dataset, an additional fully-automatic cavity filling was performed using the 3D Slicer software “wrap solidify” filter (outer surface extraction, minimum set to 10mm) [1] and mandible mask predictions were analyzed.

1. Weidert S, Andress S, Linhart C, et al (2020) 3D printing method for next-day acetabular fracture surgery using a surface filtering pipeline: feasibility and 1-year clinical results. Int J Comput Assist Radiol Surg 15:565–575. https://doi.org/10.1007/s11548-019-02110-0

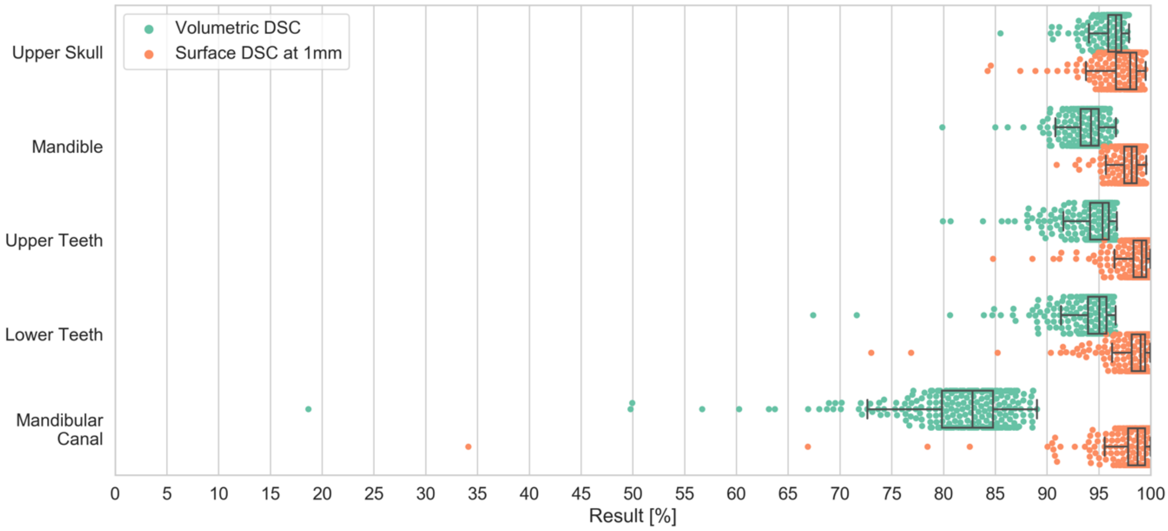


**Supplementary Figure 1.** Volumetric DSC and surface DSC at 1mm results of our 5-fold cross-validation (300 CT scans).

**Supplementary Table 1.** Mean ± Standard Deviation quantitative results of our 5-fold cross-validation (300 CT scans)

| | Upper Skull | Mandible | Upper teeth | Lower Teeth | Mandibular canal | Total |
|---|---|---|---|---|---|---|
| Volume DSC | 0.9632 ± 0.0138 | 0.9386 ± 0.0179 | 0.9455 ± 0.024 | 0.9414 ± 0.0309 | 0.814 ± 0.0656 | 0.9204 ± 0.0648 |
| Jaccard Coefficient | 0.9294 ± 0.0248 | 0.8848 ± 0.0305 | 0.8976 ± 0.0407 | 0.8907 ± 0.0495 | 0.6906 ± 0.0788 | 0.6773 ± 0.3595 |
| Volume Similarity | -0.0043 ± 0.0421 | -0.0136 ± 0.0612 | -0.0194 ± 0.0583 | -0.0188 ± 0.0683 | -0.0160 ± 0.0799 | 0.1668 ± 0.3678 |
| Average Surface Distance (GT to Prediction) | 0.1491 ± 0.098 | 0.1167 ± 0.0546 | 0.1021 ± 0.0825 | 0.1049 ± 0.0985 | 0.1985 ± 0.5738 | 0.1344 ± 0.2703 |
| Average Surface Distance (Prediction to GT) | 0.0698 ± 0.0677 | 0.0866 ± 0.0556 | 0.0909 ± 0.2332 | 0.1161 ± 0.4811 | 0.1646 ± 0.2508 | 0.1056 ± 0.2684 |
| Hausdorff Distance 100% (mm) | 9.6152 ± 3.9791 | 4.0386 ± 1.7017 | 3.9237 ± 7.6032 | 3.8274 ± 7.1537 | 3.341 ± 2.7371 | 4.9541 ± 5.6882 |
| Hausdorff Distance 95% (mm) | 0.8863 ± 0.7018 | 0.6942 ± 0.2767 | 0.5909 ± 0.3977 | 0.8066 ± 2.7852 | 0.8845 ± 1.9401 | 0.7731 ± 1.5674 |
| Surface DSC at 1mm | 0.9736 ± 0.0205 | 0.9791 ± 0.0117 | 0.9860 ± 0.0176 | 0.9832 ± 0.0262 | 0.9786 ± 0.0473 | 0.9801 ± 0.0278 |

**Supplementary Table 2.** Mean ± Standard Deviation quantitative results on our test dataset (153 CT scans)

| | Upper Skull | Mandible | Upper teeth | Lower Teeth | Mandibular canal | Total |
|---|---|---|---|---|---|---|
| Volume DSC | 0.9622 ± 0.0143 | 0.9419 ± 0.0162 | 0.9483 ± 0.0181 | 0.9438 ± 0.0232 | 0.8159 ± 0.0579 | 0.9224 ± 0.0619 |
| Jaccard Coefficient | 0.9274 ± 0.0258 | 0.8907 ± 0.0279 | 0.9022 ± 0.0316 | 0.8944 ± 0.0389 | 0.6928 ± 0.076 | 0.8615 ± 0.0961 |
| Volume Similarity | -0.0133 ± 0.0412 | -0.0118 ± 0.0571 | -0.0183 ± 0.0527 | -0.0177 ± 0.0606 | -0.0119 ± 0.0819 | -0.0146 ± 0.0601 |
| Average Surface Distance (GT to Prediction) | 0.1695 ± 0.138 | 0.1137 ± 0.0508 | 0.0912 ± 0.0537 | 0.102 ± 0.1119 | 0.1935 ± 0.2908 | 0.134 ± 0.1608 |
| Average Surface Distance (Prediction to GT) | 0.0625 ± 0.0421 | 0.0807 ± 0.0488 | 0.0972 ± 0.3492 | 0.0779 ± 0.0697 | 0.157 ± 0.1364 | 0.0951 ± 0.1754 |
| Hausdorff Distance 100% (mm) | 9.4477 ± 4.2425 | 4.0158 ± 1.5094 | 4.6572 ± 18.8037 | 3.5852 ± 2.7675 | 3.4583 ± 2.933 | 5.0328 ± 9.078 |
| Hausdorff Distance 95% (mm) | 1.0097 ± 0.8568 | 0.697 ± 0.2868 | 0.5403 ± 0.2273 | 0.589 ± 0.5276 | 0.9853 ± 1.9992 | 0.7642 ± 1.0317 |
| Surface DSC at 1mm | 0.9692 ± 0.0308 | 0.9792 ± 0.0122 | 0.9887 ± 0.0118 | 0.9853 ± 0.02 | 0.979 ± 0.0351 | 0.9803 ± 0.0248 |

**Supplementary Table 3.** Quantitative results on public mandible test dataset (10 CT scans)

| | Prediction vs Operator | Mandible number | | | | | | | | | |
|---|---|---|---|---|---|---|---|---|---|---|---|
| | | 1 | 2 | 3 | 4 | 5 | 6 | 7 | 8 | 9 | 10 |
| Volume DSC | A | 0.9172 | 0.8512 | 0.8929 | 0.8938 | 0.9216 | 0.9074 | 0.9215 | 0.8871 | 0.9266 | 0.6119 |
| | B | 0.9147 | 0.8468 | 0.8980 | 0.8911 | 0.9203 | 0.9129 | 0.9277 | 0.8823 | 0.9257 | 0.6146 |
| Jaccard Coefficient | A | 0.8471 | 0.7409 | 0.8065 | 0.8079 | 0.8545 | 0.8306 | 0.8544 | 0.7971 | 0.8632 | 0.4408 |
| | B | 0.8428 | 0.7344 | 0.8149 | 0.8037 | 0.8524 | 0.8398 | 0.8652 | 0.7894 | 0.8617 | 0.4437 |
| Volume Similarity | A | 0.0410 | 0.0200 | 0.0325 | 0.0175 | 0.0200 | -0.0135 | 0.0070 | 0.1420 | -0.0234 | 0.6886 |
| | B | 0.0041 | 0.0411 | 0.0720 | 0.0370 | 0.0185 | -0.0013 | -0.0072 | 0.1641 | -0.0315 | 0.6802 |
| Average Surface Distance (GT to Prediction) | A | 0.2620 | 0.4735 | 0.2562 | 0.3892 | 0.2329 | 0.2820 | 0.2834 | 0.4199 | 0.2221 | 26.8738 |
| | B | 0.2644 | 0.4933 | 0.2382 | 0.4205 | 0.2385 | 0.2593 | 0.2573 | 0.4463 | 0.2256 | 27.0208 |
| Average Surface Distance (Prediction to GT) | A | 0.2201 | 0.2138 | 0.1908 | 0.1846 | 0.1721 | 0.2256 | 0.1557 | 0.2437 | 0.1789 | 0.2053 |
| | B | 0.2302 | 0.2277 | 0.1717 | 0.1972 | 0.1753 | 0.2176 | 0.1687 | 0.2591 | 0.1775 | 0.2063 |
| Hausdorff Distance 100% (mm) | A | 4.0352 | 11.3262 | 4.0872 | 5.1444 | 2.5488 | 2.8038 | 3.8194 | 6.9552 | 2.9770 | 90.3069 |
| | B | 4.0000 | 10.9110 | 3.1053 | 4.2960 | 2.5993 | 3.0000 | 3.7950 | 6.9552 | 2.9770 | 90.4536 |
| Hausdorff Distance 95% (mm) | A | 1.0000 | 2.0000 | 1.2598 | 1.9994 | 0.7209 | 1.3037 | 1.0000 | 2.0000 | 0.7500 | 80.9020 |
| | B | 1.0000 | 2.0615 | 0.9390 | 1.9994 | 1.0000 | 1.0307 | 1.0000 | 2.0000 | 0.7500 | 81.0128 |
| Surface DSC at 1mm | A | 0.9710 | 0.9296 | 0.9562 | 0.9056 | 0.9803 | 0.9412 | 0.9729 | 0.9084 | 0.9910 | 0.6386 |
| | B | 0.9708 | 0.9232 | 0.9636 | 0.8952 | 0.9775 | 0.9481 | 0.9801 | 0.9002 | 0.9903 | 0.6404 |
